# EasyCOV : LAMP based rapid detection of SARS-CoV-2 in saliva

**DOI:** 10.1101/2020.05.30.20117291

**Authors:** Nicolas L’Helgouach, Pierre Champigneux, Francisco Santos Schneider, Laurence Molina, Julien Espeut, Mellis Alali, Julie Baptiste, Lise Cardeur, Benjamin Dubuc, Vincent Foulongne, Florence Galtier, Alain Makinson, Grégory Marin, Marie-Christine Picot, Alexandra Prieux-Lejeune, Marine Quenot, Francisco Checa Robles, Nicolas Salvetat, Diana Vetter, Jacques Reynes, Franck Molina

**Affiliations:** Sys2Diag UMR9005 CNRS / ALCEDIAG, Montpellier, France; Virology department, Montpellier University Hospital, France; INSERM Centre Investigation Clinique 1411, University Hospital, Montpellier, France; Infectious diseases department, University Hospital, Montpellier, France; DIM Clinical Research Unit, University Hospital, Montpellier, France

**Author notes:** Contribute equally to the work (Nicolas L’Helgouach, Pierre Champigneux). Contribute equally to the work (Jacques Reynes, Franck Molina). Corresponding authors: Franck Molina, Sys2diag CNRS.

## Abstract

Covid-19 crisis showed us that rapid massive virus detection campaign is a key element in SARS-CoV-2 pandemic response. The classical RT-PCR laboratory platforms must be complemented with rapid and simplified technologies to enhance efficiency of large testing strategies.

To this aim, we developed EasyCOV, a direct saliva RT-LAMP based SARS-CoV-2 virus detection assay that do not requires any RNA extraction step. It allows robust and rapid response under safe and easy conditions for healthcare workers and patients.

EasyCOV test was assessed under double blind clinical conditions (93 asymptomatic healthcare worker volonteers, 10 actively infected patients, 20 former infected patients tested during late control visit). EasyCOV results were compared with classical laboratory RT-PCR performed on nasopharyngeal samples.

Our results show that compared with nasopharyngeal laboratory RT-PCR, EasyCOV SARS-CoV-2 detection test has a sensitivity of 72.7%. Measured on healthcare worker population the specificity was 95.7%. LAMP technology on saliva is clearly able to identify subjects with infectivity profile. Among healthcare worker population EasyCOV test detected one presymptomatic subject.

Because it is simple, rapid and painless for patients, EasyCOV saliva SARS-Cov-2 detection test may be useful for large screening of general population.

## Introduction

COVID-19 crisis teaches us that before, during and after population lock-down, massive testing is a relevant strategy to protect worldwide populations. However, to be efficient, testing strategy must be compliant with a vast variety of use cases ranging from sanitary to socio-economics concerns. Many use cases cannot afford multi-step testing procedures including sample collection, transport, laboratory analysis, data treatment and patient information leading to at least 12h to 24h response time. This response delay induces efficiency loss in all testing strategies. In addition, some use cases require quick answers on the spot like airport, restricted area access, crowded close zone, etc. In such cases, virus detection assays must be quickly and easily performed at low cost in a minimally invasive way.

Current SARS-CoV-2 Virus detection assays are mainly of two kinds: i) antigen assays based on viral antigen detection using immunoassays technology or ii) RT-PCR based approaches. Laboratory RT-PCR approaches are clearly the most sensitive (and specific) however their real clinical performances may be attenuated by the sampling procedure (X. Li et al., 2020). Nasopharyngeal (NP) sampling is the most widely used. NP laboratory RT PCR sensitivity is around 70%. In addition to the necessity of the NP sampling to be performed by qualified medical staff, it is painful for the patient and at risk to contaminate the caregiver (Wen-Liang Guo et al., 2020).

Recent publications showed that Saliva is a reliable tool to detect SARS-CoV-2 (Azzi et al., 2020) with strong virus load in contagious patients. In addition, some studies suggested that saliva is more sensitive to SARS-CoV-2 with higher titers than NP swabs (Wyllie et al., 2020). The fact that SARS-CoV-2 virus was found to have salivary gland tropism in animal may be an explanation (Jasper Fuk-Woo Chan et al., 2020). Comparative studies in human show that SARS-CoV-2 virus content in saliva may appear sooner than in NP swabs. Adding the fact that saliva sampling procedure is simpler for caregiver and less invasive for patient makes saliva sample of great interest in the context of regular on the spot testing strategy (Kelvin Kai-Wang To et al., 2020). In addition the consistency seems to be higher in saliva throughout the course of infection (Willie et al., 2020).

On the technological point of view, RT-LAMP isothermal amplification technology is very attractive since it allows rapid RNA reverse transcription and DNA amplification without temperature cycles requirement. A simple 65°C heating allows the detection of specific RNA fragments. It has already been used for SARS-CoV-2 specific detection (Anhatar et al., 2020, Tsai et al., 2020, Ben-Assa et al., 2020, Thi al., 2020, Minghua Jiang et al, 2020; Yinhua Zhang et al., 2020; Laura E. Lamb et al., 2020). LAMP technology does not even require complex devices neither laboratory infrastructures. One limiting step of the actuals laboratory RT-PCR SARS-CoV-2 detections is the RNA extraction step that is time consuming, expensive and require centrifugation steps.

We developed a direct saliva assays avoiding RNA extraction (EasyCOV test). For that, we had to tackle human saliva complexity and variability. We compared herein EasyCOV, a raw salivary SARS-CoV-2 virus assay to NP RT-PCR SARS-CoV-2 patient assessment. It has been designed to ease the sampling, protect the caregivers and to return a reliable rapid response at low cost. Thus, it can be used as on the spot security net in the context of Covid-19 crisis.

## Materials and Methods

### Clinical Samples

Prior to nasopharyngeal swabbing for SARS-CoV-2 RT-PCR, healthcare workers were interviewed for symptoms on the day and during the 10 past days. For patients, these symptoms were collected in the medical record as well as respiratory complications and specific CT scan signs.

Saliva samplings were collected at the same time as nasopharyngeal swabbing for all healthcare workers having given their agreement to participate in the study. For patients already tested, only the saliva samples have been redone. The results of RT-PCR already carried out were all collected in the electronic CRF of the study. All the nasopharyngeal RT-PCR were analyzed in the Montpellier University Hospital Virology laboratory while saliva sampling were sent to research laboratory Sys2Diag (Figure 1).

**Figure 1.**
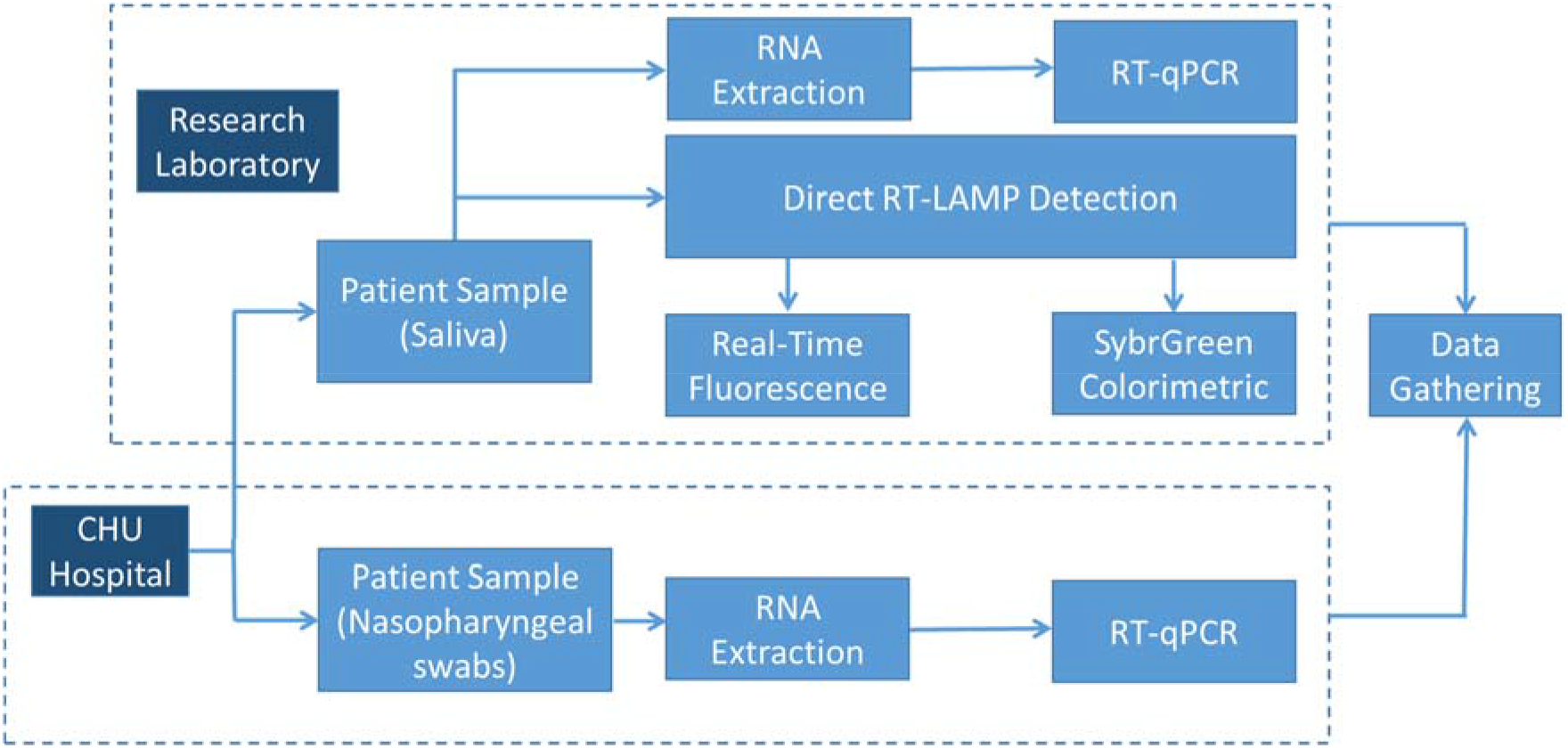
Clinical study work-flow. Samples available for this study were taken by the hospital. Saliva samples were put on ice and transferred to the laboratory within 3 hours. RNA from nasopharyngeal samples were extracted and submitted to SARS-CoV-2 detection by RT-PCR at the hospital. Saliva samples were submitted to different tests in the research laboratory: (i) RNA extraction and SARS-CoV- 2 detection by RT-PCR, (ii) Direct SARS-CoV-2 detection without RNA extraction by RT-LAMP combined to Fluorescence and colorimetric reading in parallel.

### RNA extraction from nasopharyngeal sampling and RT-qPCR SARS-CoV-2 virus detection

Samples were collected through nasopharyngeal swabbing further discharged in 1 mL viral transport medium (VTM). Prior to RNA extraction, 200 µL of VTM were inactivated by mixing with the same amount of ATL Lysis buffer (Qiagen, Germany). Extraction was performed on 200 µL of inactivated samples using an automatic extraction process running the Starmag extraction kit (Seegene, Korea). A commercial multiplexed real time RT-PCR targeting the RdRp, E and N viral genes was used for viral detection (Allplex 2019-nCov assay kit, Seegene, Korea).

### Saliva pretreatment and direct LAMP SARS-CoV-2 virus detection

In order to inactivate the RT-LAMP inhibitory components in the saliva, the saliva were incubated at 65°C for 30 minutes before RT-LAMP reaction.

After treatment, 3 µl of treated saliva were added to a RT-LAMP reaction mix containing 1X Warmstart buffer (NEB E1700L), 1M Betaine and a DNA-primer mix composed of 1,6 µM FIP, 1,6 µM BIP, 0,2 µM F3, 0,2 µM B3, 0,4 µM LOOP F (F1), 0,4 µM LOOP B (B1). For real-time detection on a real-time PCR system (Roche LightCycler 480), we added to the reaction a fluorescent dye (NEB E1700L). For EasyCOV colorimetric detection, after 30 minutes of incubation at 65°C we added 1µl of Sybrgreen I (Invitrogen S7563). Saliva positive for SARS-CoV-2 turned yellow while negative ones were orange. The final color can be automatically interpreted by a smartphone application called EasyCOV Reader developed by VOGO Company. For each experiment we added positive and negative controls: a SARS-CoV-2 purified RNA (Human 2019-nCoV strain 2019-nCoV/Italy-INMI1 RNA #008N-03894 - EVAg), 1 confirmed positive saliva, 1 confirmed negative saliva, a No template control (H2O) and a SARS DNA (Not cross-reacting with SARS-CoV-2, IDT, #10006624).

### RNA extraction from saliva and RT-qPCR SARS-Cov-2 virus detection (Based on Pasteur Institute protocol on nasopharyngeal sampling)

After collection, 100 µl of saliva samples were treated in the presence of 5mM DTT for 30 minutes at RT. After treatment, saliva RNA extraction was done using the Nucleospin Dx Virus Kit (Macherey Nagel #740895.50). RT-qPCR reactions were prepared in 384 multi-well plates. After purification, 2 µl of purified RNA were added to a Invitrogen superscript III Platinium One step reaction mix (#11732020) containing 1X reaction mix, 0,8 mM MgSO4 and a DNA-primer mix composed of 0,4 µM forward primer nCoV_IP2-12669, 0,4 µM reverse primer IP2-12759, 0,4 µM forward primer nCoV_IP4-14059, 0,4 µM reverse primer nCoV_IP4-14146, 0,16 µM nCoV IP2-12696b Probe(+), 0,16 µM nCoV_IP4-14084 Probe(+). Real-time detection was performed on a real-time PCR system (Roche LightCycler 480) directed by the LightCycler 480 Software (version 1.5.1.62). Thermal cycling conditions were: Reverse transcription 20 minutes at 55°C, pre-Incubation 3 minutes at 95°C, 50 cycles of DNA amplification (95°C for 15s, 58°C for 30s) and 30s of cooling at 40°C.

### Statistical Analysis

All statistics and figures were performed with the “R” statistical open source Software (version 3.5.3). Contingency table and diagnostic performance was calculated using the confusion Matrix function in the caret R package version 6.0-86 (Kuhn, 2008).

## Results and Discussion

### Design of Study

In this monocentric prospective diagnostic test study, patients or subjects were prospectively included. The salivary SARS-CoV-2 virus assay (EasyCOV) was analyzed double-blinded on COVID-19 status and on reference test (RT-PCR on nasopharyngeal sampling).

Salivary clinical samples were recovered from patients with a previous known SARS-COV-2 infection admitted to University Montpellier Hospital in the unit specialized in COVID and from volunteer caregivers working at the same hospital. Healthcare workers or adult outpatients were recruited when attending the center for SARS-CoV-2 screening or diagnosis, based on symptomatic case-finding or close exposure with an index case.

For this intermediary analysis, a total of 130 subjects were recruited (Figure 2). Among them, 7 subjects were excluded from the analysis due to the insufficient quantity of salivary material for testing. This clinical evaluation was approved by French National Protection of Person Committee (NCT04337424).

**Figure 2.**
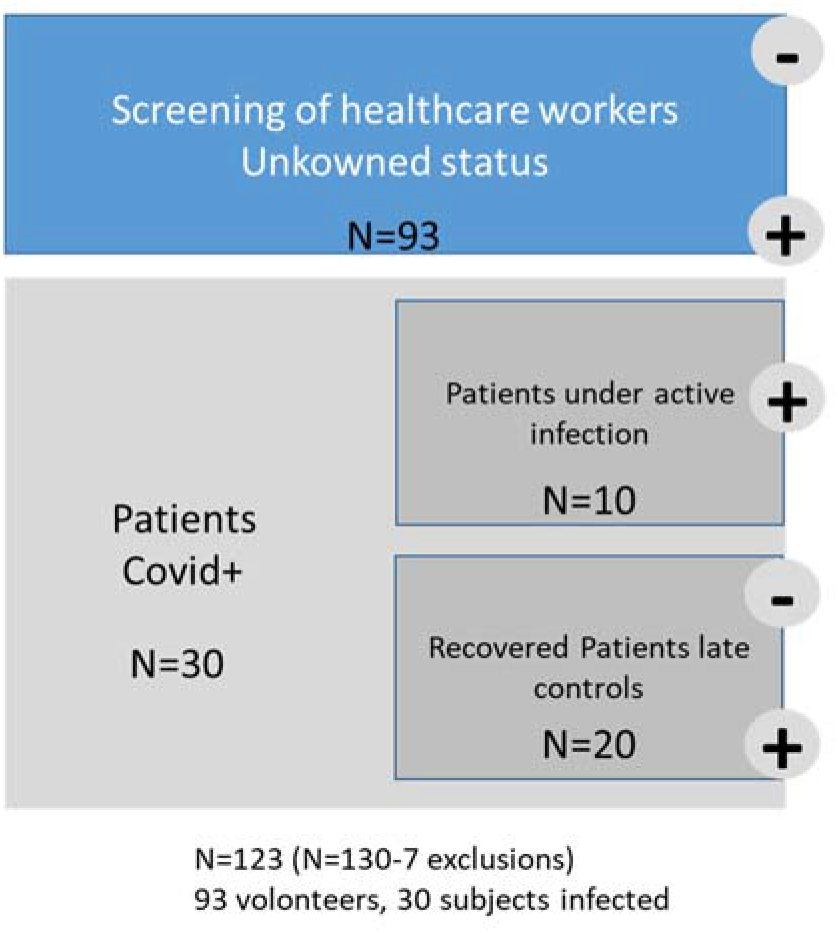
Subject stratification.

Screening of healthcare workers of unknown Covid status were categorized separately from patients Covid+. Patients Covid+ were separated in two classes: One corresponding to patients under active infection (Ct< 35), a second class correspond to patients who recovered from Covid-19 (Ct> = 35) and inspected for a late control visit. It was demonstrated by La Scola et al that infectivity disappears when Ct measured with NP RT-PCR was higher that 34 (La Scolla et al., 2020).

Considering that covid-19 was more severe for aged population, as expected, the average age of active patients is higher than healthcare worker population. The same related with the inflammation markers (e.g. CRP) was observed. Serology was only performed on healthcare workers.

## EasyCOV a simple and quick SARS-CoV-2 saliva detection for mass screening

Here we developed a new simple saliva SARS-CoV-2 detection test based on RT-LAMP technology that we called EasyCOV (Figure 3). After saliva collection with a pipette under the tongue, 200 µl of saliva are transferred in a 0.5 ml tube in which a salivary inactivation solution is present. The tube is then incubated at 65°C for 30 minutes in a simple thermoblock. After incubation, 3 µl of inactivated saliva is transferred in a new tube containing 17 µl of RT-LAMP reaction mix. After 30 minutes of incubation at 65°C, the RT-LAMP reaction is revealed by the addition of 1µl of SybrGreen. A yellow coloration of the mix gives a positive result for the presence of the virus. An orange coloration gives a negative result. After addition of the SybrGreen, to get a precise reading, the tube is analyzed thanks to a specific mobile application (EasyCOV Reader, VOGO) installed on a mobile phone or a touch pad. In order to keep the detection usable with different amount and quality of light, the detection is done with the tube laying on a white paper were colors of reference are printed (Figure 4). The mobile Apps will give three possible responses: Positive, Negative or Ambiguous.

**Figure 3.**
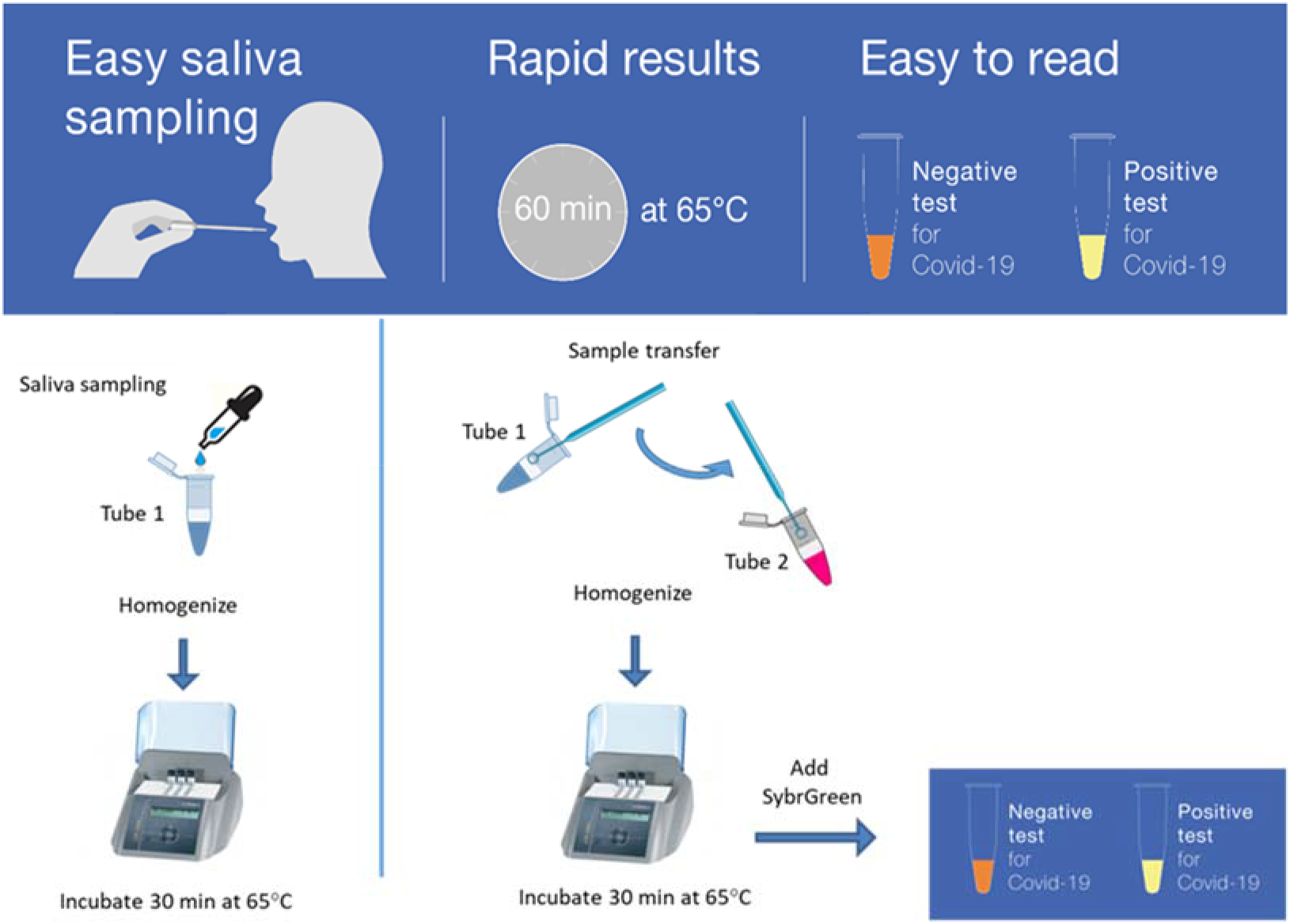
Schematic representation of EasyCOV steps for SARS-CoV-2 detection

**Figure 4.**
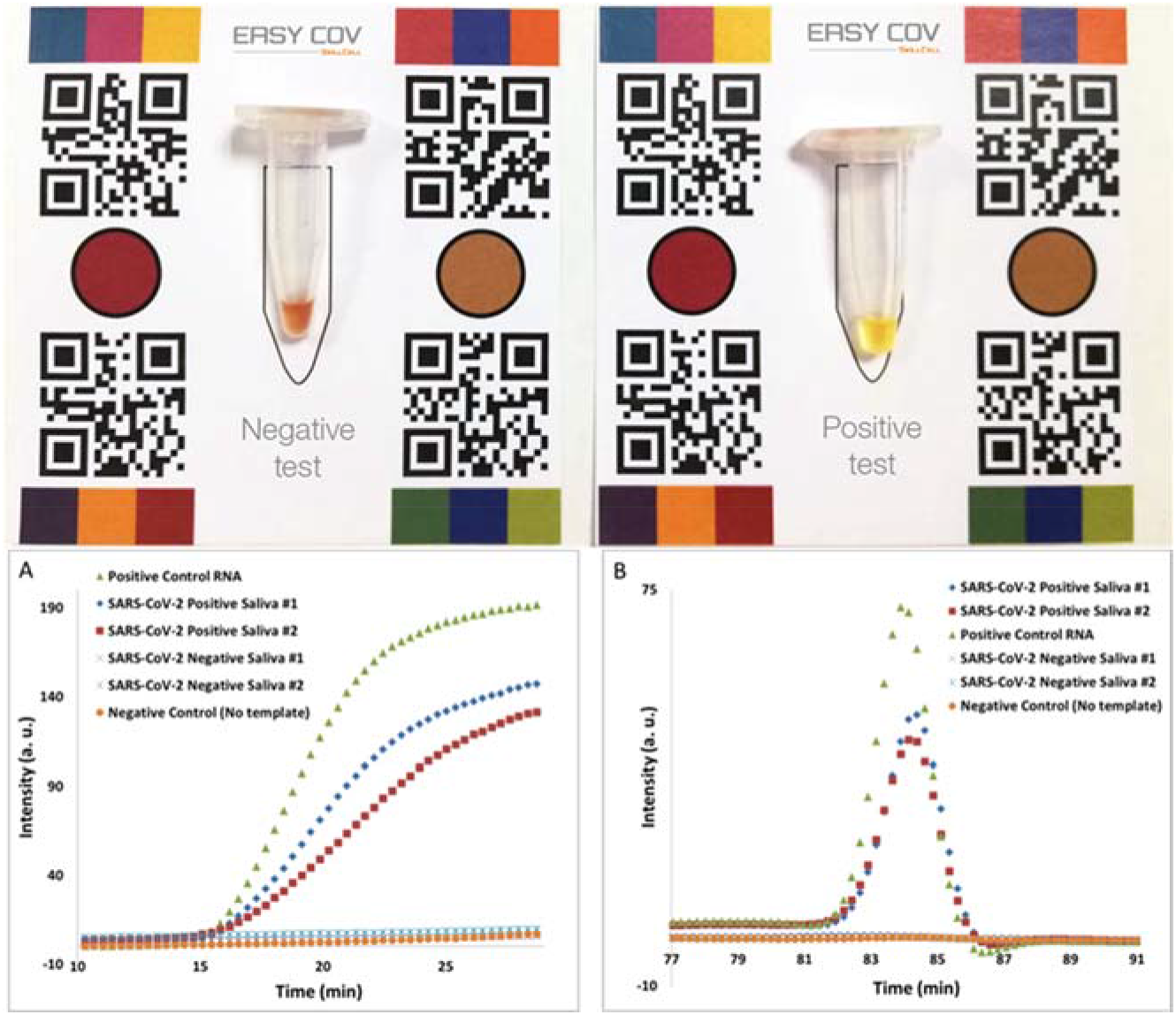
EasyCOV colorimetric assay and fluorescence real-time analysis of RT-LAMP amplification of SARS-CoV-2 virus. Upper figure: EasyCOV colorimetric assay. Readout of a positive and a negative patient saliva. Lower figure: (A) Representative fluorescence signals obtained for RT-LAMP amplification of 2 positively and 2 negatively infected saliva. A positive control containing SARS-CoV-2 purified RNA and a negative control lacking template has been included. (B) Representative melting curves analysis of the samples.

## EasyCOV Saliva test evaluation

In order to evaluate our EasyCOV RT-LAMP assay, we compared it to the Montpellier University Hospital clinical status based on NP RT-PCR Ct threshold (< 35 is considered as positive and potentially infectious) (Figure 1, Table S1 and Table S2). To follow the RT-LAMP amplification process we performed in parallel end-point colorimetric experiment (similar to real EasyCOV assay) and real-time fluorescence tracing of RT-LAMP amplification followed by Tm analysis (Figure 4). Each experiment was run with positive and negative controls. The lamp Tm analysis was stated according to control profile. This fluorescence parallel study was a way to monitor the EasyCOV RT-LAMP assay.

All the sample collected from all the groups were analyzed using EasyCOV RT-LAMP assay and compared against reference NP RT-PCR performed in double blinded at Montpellier University Hospital and Sys2diag laboratory. The results were expressed as qualitative patient positive/negative status and included in the contingency table 2. Most of the negative samples found by the Montpellier University Hospital were found negative using EasyCOV test. Four false positive were found in the group healthcare workers. One False positive found with EasyCOV belong to the group of covid-19 recovered patients during their late control visit. Consequently the specificity of EasyCOV for heathlcare workers screening was 95.7%.

**Table 1.**
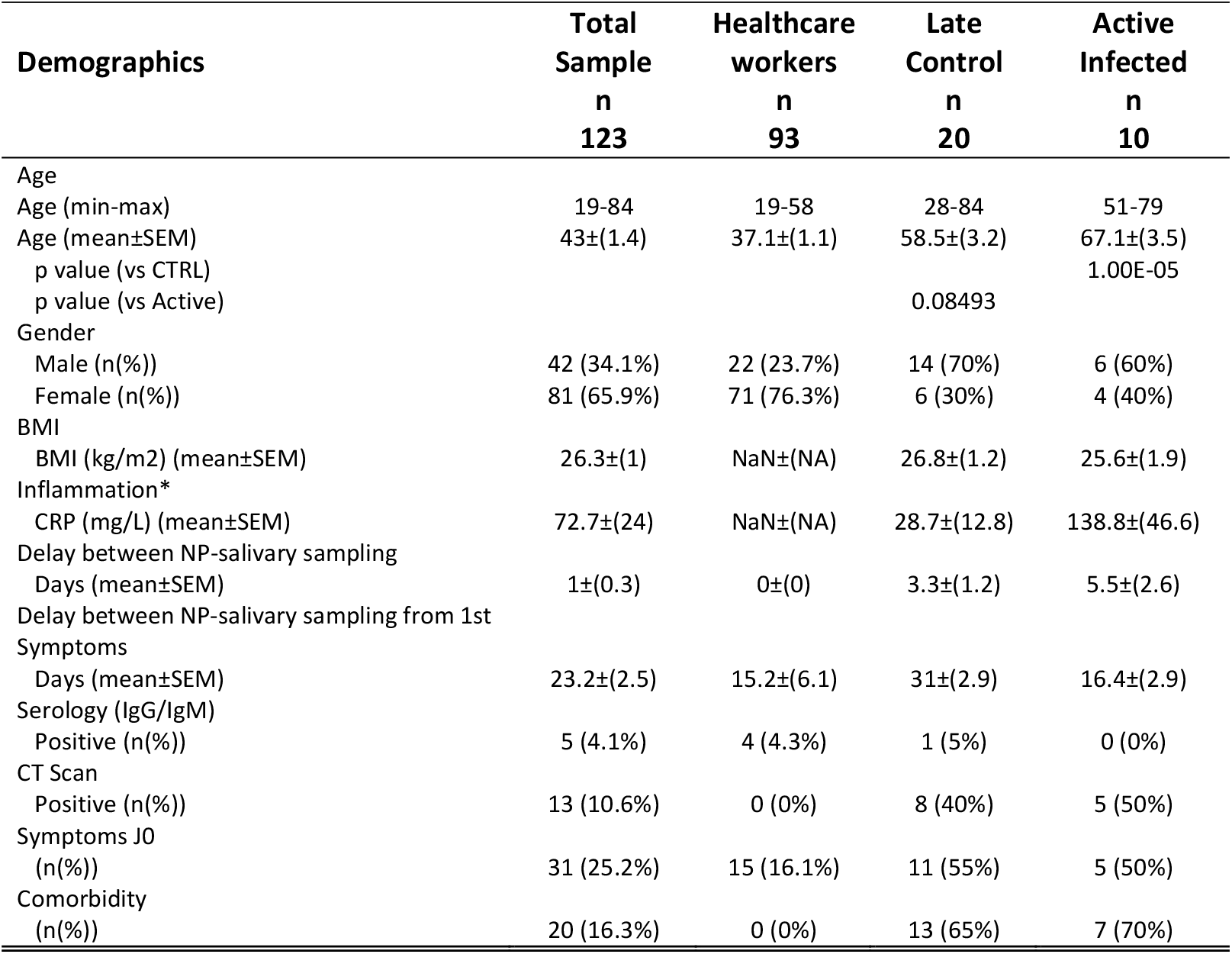
Clinical characteristic of cohort

**Table 2.** Contingency table of the results shown in table S1 and S2, comparing subject status based on nasopharyngeal RT-qPCR and salivary EasyCOV test for 123 subjects. (Right) Sensitivity and specificity of the EasyCOV test with respect to the subject status.

|  |  | Subject status based on<br>RT-qPCR |  |  |  | Total |
| --- | --- | --- | --- | --- | --- | --- |
|  |  | Heathcare<br>workers | Active<br>Infection | Late<br>control |  |  |
|  |  | Neg | Pos | Neg | Pos |  |
| Easycov<br>Results | Neg | 88 | 3 | 11 | 6 | 108 |
|  | Pos | 4 | 8 | 1 | 2 | 15 |
|  | Total | 92 | 11 | 12 | 8 | 123 |

|  | Sensitivity | 95 % CI |
| --- | --- | --- |
| Active Infection (ct <35) | 72.7% | [39.0% ; 94.0%] |
| Positive late control (Ct ≥35) | 25.0% | [3.2% ; 65.1%] |

|  | Specificity | 95 % CI |
| --- | --- | --- |
| Negative Healthcare workers | 95.7% | [89.2% ; 98.8%] |
| Negative Late control | 91.7% | [61.5% ; 99.8%] |

Among the patients under active infection (NP RT-PCR Ct< 35) we observed a sensitivity of 72.7% with EasyCOV test. This makes EasyCOV test suitable for on the spot SARS-CoV-2 population screening.

Our study represent a typical distribution of population for a screening campaign with a large group of non-infected people, and a population with either a recent or late infection. In this context EasyCOV is able to detect about 73% of infected patients. However, specific cases have been identified as representative of typical profiles that can be encounter under real conditions.

Infectious patients like patient P#010 are likely to be detected, although a few of them could be missed. As well negative subjects like S#018 have been stated negative by EasyCOV and a very low amount of false positive was observed. Next to these simple cases other patients highlight less obvious cases that may occur when use at large scale.

Presymptomatic patients are always difficult cases to address since their viral content may strongly vary before the first symptoms. For instance, two days before the first symptoms the viral content is supposed to be very high however they are big discrepancies from one patient to another. In our study, we had the case of P#080 who was found negative with NP RT PCR but positive using EasyCOV in saliva. For this patient, investigation with RT PCR from RNA extraction from saliva was found positive with Ct = 28.8. In addition, this patient was seropositive for SARS-CoV-2. Thus, EasyCOV, in this case, was able to identify a presymptomatic patient.

Few false positive results were raised by EasyCOV. When analyzed, we saw two kinds of false positive. In the first kind are the patients that have never been infected with all other virological and serological tests negative but found positive with LAMP in saliva. They are probably real false positive. The second category, are the patients that have been infected, with at least one virological or serological test positive but are considered as cured at the time of saliva sampling. In such case, some patients were still found positive by EasyCOV, probably signing low content of active or inactive virus. If the serology is positive, one can say that EasyCOV test detects patients with late infection. On the other hand, if the serology is negative, then the patient may be under active infection and thus, must be controlled by a nasopharyngeal RT-PCR assay.

## Conclusion

EasyCOV SARS-CoV-2 virus detection test was designed in the context of covid-19 response. Because EasyCOV is a rapid and simple, it is compatible with large scale population screening. In addition, saliva who was shown to be reliable sample for SARS-CoV-2 virus detection, is less invasive for tested subject and thus prone to be easily accepted by the population.

This clinical study was perform on distinct subject groups. Healthcare workers with unknown covid status and covid+ patients. This later group was divided in two subgroups patients actively infected (NP RT-qPCR Ct< 35) and cured patients on late visit (NP RT-qPCR Ct> = 35). These groups were supposed to represent a classical screening population distribution.

EasyCOV performances was 72.7% sensitivity on actively infected patients. But was still able in some case to detect virus in cured patients on late control visit. The specificity was 95.7% meaning a very low rate of false positive result. However, among healthcare worker population EasyCOV test detected one presymptomatic subject. Future works must investigate the real status of the remaining few false positive.

EasyCOV saliva SARS-CoV-2 detection is then suitable for on the spot use in the context of population screening.

## Data Availability

All data from this study are available on demand.

## Ethics and Reporting

The intend of this work was to develop clinically compliant rapid virus detection test in the context of emergency concerns of covid-10 crisis. The work was undertaken in accordance with the French ethical requirements. The clinical study was reviewed (CPP Comity of Person Protection) accepted under the registration number 20036-60435. Individual consent is a part of the sample collection procedure. Human 2019-nCoV strain 2019-nCoV/Italy-INMI1 RNA (#008N-03894) was furnished by EVAg company under a biological material transfer agreement (Horizon 2020 programme Grant No. 653316).

Reference of Clinical assay: NCT04337424

## Acknowledgements

We thanks Nicolas Doll from New England Biolabs France for technical support, Caroline Goujon CNRS for helpful scientific discussion, Victor Petit, Sofia El Annabi, Alexandre Lassaigne from Skillcell, Patrick Collet, Yann Pichot, Marine Bittel from TRONICO, Marc Delmas, from PMB, Pascaline Dubs, Guillaume Rochet from CNRS, Thomas Hottier and Jérome Bayette from Labosud, Christophe Carniel from VOGO for technical discussions, developments and kind support. Sys2diag members for their day basis support during such complicated period.

This work was financially supported by the CNRS, SkillCell, Region Occitanie and DGA (AID).

---

**Table S1.** Clinical profiles and virus detection from patient group.

|  |  |  | NP RT-qPCR<br>(Ct) | Salivary<br>RT-LAMP | Delay between<br>NP - salivary<br>sampling | Delay between<br>first symptoms and<br>salivary sampling | Symptoms at the<br>day of salivary<br>sampling | Clinical Status |
| --- | --- | --- | --- | --- | --- | --- | --- | --- |
| Subjects with a previous known infection | Subjects Currently<br>infected (Ct < 35) | P007 | 18 | NEG | 7 | 10 | Y | Infected |
|  |  | P010 | 25 | POS | 6 | 14 | N | Infected |
|  |  | P011 | 33 | POS | 5 | 16 | N | Infected |
|  |  | P013 | 32 | NEG | 2 | 24 | Y | Infected |
|  |  | P014 | 33 | POS | 4 | 24 | Y | Infected |
|  |  | P015 | 29 | POS | 0 | 21 | Y | Infected |
|  |  | P016 | 32 | POS | 0 | 3 | Y | Infected |
|  |  | P017 | 21 | NEG | 3 | 7 | N | Infected |
|  |  | P118 | 25 | POS | 28 | 29 | N | Infected |
|  |  | S129 | 34 | POS | 0 | - | N | Infected |
|  | Late Control (Ct ≥ 35) | P004 | 38 | NEG | 2 | 27 | N | Recovered/Non-Infected |
|  |  | P006 | 39 | NEG | 9 | - | N | Recovered/Non-Infected |
|  |  | P008 | 37 | NEG | 4 | 8 | N | Recovered/Non-Infected |
|  |  | P012 | 38 | NEG | 0 | - | N | Recovered/Non-Infected |
|  |  | P027 | 40 | POS | 0 | 31 | Y | Recovered/Non-Infected |
|  |  | P055 | 36 | NEG | 2 | 15 | N | Recovered/Non-Infected |
|  |  | S122 | 39 | POS | 0 | 23 | Y | Recovered/Non-Infected |
|  |  | P127 | 37 | NEG | 5 | 29 | N | Recovered/Non-Infected |
|  | Late Control (Negative) | P005 | NEG | NEG | 0 | 21 | N | Recovered/Non-Infected |
|  |  | P074 | NEG | NEG | 0 | 37 | Y | Recovered/Non-Infected |
|  |  | P075 | NEG | NEG | 14 | 36 | N | Recovered/Non-Infected |
|  |  | P097 | NEG | NEG | 0 | 37 | N | Recovered/Non-Infected |
|  |  | P114 | NEG | NEG | 0 | 41 | N | Recovered/Non-Infected |
|  |  | P115 | NEG | NEG | 0 | - | N | Recovered/Non-Infected |
|  |  | P117 | NEG | NEG | 0 | 35 | N | Recovered/Non-Infected |
|  |  | S119 | NEG | NEG | 0 | 54 | Y | Recovered/Non-Infected |
|  |  | P120 | NEG | NEG | 17 | 39 | Y | Recovered/Non-Infected |
|  |  | S121 | NEG | NEG | 0 | 21 | Y | Recovered/Non-Infected |
|  |  | P126 | NEG | NEG | 13 | 42 | Y | Recovered/Non-Infected |
|  |  | S130 | NEG | POS | 0 | - | N | Recovered/Non-Infected |

NEG= Negative result. POS= Positive result. Y = Presence of at least one symptom related to COVID-19 disease. N= Absence of symptoms related to COVID-19 disease. (-) = Not determined.

**Table S2.** Clinical profiles and virus detection from healthcare workers group.

|  |  | NP RT-qPCR<br>(Ct) | Salivary<br>RT-LAMP | Delay between<br>NP - salivary<br>sampling | Delay between first<br>symptoms and<br>salivary sampling | Symptoms at<br>the day of<br>salivary<br>sampling | Clinical Status |
| --- | --- | --- | --- | --- | --- | --- | --- |
| Volunteers health care workers | S009 | NEG | NEG | 0 | - | N | Recovered/Non-Infected |
|  | S018 | NEG | NEG | 0 | - | N | Recovered/Non-Infected |
|  | S020 | NEG | NEG | 0 | - | N | Recovered/Non-Infected |
|  | S021 | NEG | NEG | 0 | - | N | Recovered/Non-Infected |
|  | S022 | NEG | NEG | 0 | - | Y | Recovered/Non-Infected |
|  | S023 | NEG | NEG | 0 | - | N | Recovered/Non-Infected |
|  | S024 | NEG | NEG | 0 | - | N | Recovered/Non-Infected |
|  | S025 | NEG | NEG | 0 | - | N | Recovered/Non-Infected |
|  | S026 | NEG | NEG | 0 | - | N | Recovered/Non-Infected |
|  | S028 | NEG | NEG | 0 | - | N | Recovered/Non-Infected |
|  | S030 | NEG | NEG | 0 | - | N | Recovered/Non-Infected |
|  | S031 | NEG | NEG | 0 | 36 | Y | Recovered/Non-Infected |
|  | S032 | NEG | NEG | 0 | 11 | Y | Recovered/Non-Infected |
|  | S033 | NEG | NEG | 0 | - | N | Recovered/Non-Infected |
|  | S034 | NEG | NEG | 0 | - | N | Recovered/Non-Infected |
|  | S035 | NEG | NEG | 0 | - | N | Recovered/Non-Infected |
|  | S036 | 39 | NEG | 0 | - | N | Recovered/Non-Infected |
|  | S037 | NEG | NEG | 0 | - | N | Recovered/Non-Infected |
|  | S038 | NEG | NEG | 0 | - | Y | Recovered/Non-Infected |
|  | S039 | NEG | NEG | 0 | - | N | Recovered/Non-Infected |
|  | S040 | NEG | NEG | 0 | - | N | Recovered/Non-Infected |
|  | S041 | NEG | NEG | 0 | 2 | Y | Recovered/Non-Infected |
|  | S042 | NEG | NEG | 0 | - | N | Recovered/Non-Infected |
|  | S043 | NEG | NEG | 0 | - | N | Recovered/Non-Infected |
|  | S044 | NEG | NEG | 0 | - | N | Recovered/Non-Infected |
|  | S045 | NEG | NEG | 0 | - | N | Recovered/Non-Infected |
|  | S046 | NEG | POS | 0 | - | N | Recovered/Non-Infected |
|  | S047 | NEG | NEG | 0 | - | N | Recovered/Non-Infected |
|  | S048 | NEG | NEG | 0 | - | N | Recovered/Non-Infected |
|  | S049 | NEG | NEG | 0 | - | N | Recovered/Non-Infected |
|  | S050 | NEG | NEG | 0 | - | N | Recovered/Non-Infected |
|  | S051 | NEG | NEG | 0 | - | N | Recovered/Non-Infected |
|  | S052 | NEG | NEG | 0 | 6 | Y | Recovered/Non-Infected |
|  | S053 | NEG | NEG | 0 | - | N | Recovered/Non-Infected |
|  | S054 | NEG | NEG | 0 | - | N | Recovered/Non-Infected |
|  | S057 | NEG | NEG | 0 | - | Y | Recovered/Non-Infected |
|  | S058 | NEG | NEG | 0 | - | N | Recovered/Non-Infected |
|  | S059 | NEG | NEG | 0 | - | N | Recovered/Non-Infected |
|  | S060 | NEG | NEG | 0 | - | N | Recovered/Non-Infected |
|  | S061 | NEG | NEG | 0 | 12 | Y | Recovered/Non-Infected |
|  | S062 | NEG | NEG | 0 | - | N | Recovered/Non-Infected |
|  | S063 | NEG | NEG | 0 | - | N | Recovered/Non-Infected |
|  | S064 | NEG | NEG | 0 | - | N | Recovered/Non-Infected |
|  | S065 | NEG | NEG | 0 | - | N | Recovered/Non-Infected |
|  | S066 | NEG | NEG | 0 | - | N | Recovered/Non-Infected |
|  | S067 | NEG | NEG | 0 | - | N | Recovered/Non-Infected |
|  | S068 | NEG | NEG | 0 | - | N | Recovered/Non-Infected |
|  | S069 | NEG | NEG | 0 | - | N | Recovered/Non-Infected |
|  | S070 | NEG | NEG | 0 | - | Y | Recovered/Non-Infected |
|  | S071 | NEG | NEG | 0 | - | N | Recovered/Non-Infected |
|  | S072 | NEG | NEG | 0 | - | N | Recovered/Non-Infected |
|  | S073 | NEG | NEG | 0 | - | N | Recovered/Non-Infected |
| Volunteers | S076 | NEG | NEG | 0 | - | N | Recovered/Non-Infected |
|  | S077 | NEG | NEG | 0 | - | N | Recovered/Non-Infected |
|  | S078 | NEG | NEG | 0 | - | N | Recovered/Non-Infected |
|  | S079 | NEG | NEG | 0 | - | N | Recovered/Non-Infected |
|  | S080 | NEG | POS | 0 | - | N | Recovered/Non-Infected |
|  | S081 | NEG | NEG | 0 | 2 | N | Recovered/Non-Infected |
|  | S082 | NEG | NEG | 0 | - | N | Recovered/Non-Infected |
|  | S083 | NEG | NEG | 0 | - | N | Recovered/Non-Infected |
|  | S084 | NEG | NEG | 0 | - | Y | Recovered/Non-Infected |
|  | S085 | NEG | NEG | 0 | - | N | Recovered/Non-Infected |
|  | S086 | NEG | NEG | 0 | - | Y | Recovered/Non-Infected |
|  | S087 | NEG | NEG | 0 | - | N | Recovered/Non-Infected |
|  | S088 | NEG | NEG | 0 | - | N | Recovered/Non-Infected |
|  | S089 | NEG | NEG | 0 | - | N | Recovered/Non-Infected |
|  | S090 | NEG | NEG | 0 | - | N | Recovered/Non-Infected |
|  | S091 | NEG | NEG | 0 | - | N | Recovered/Non-Infected |
|  | S092 | NEG | NEG | 0 | - | N | Recovered/Non-Infected |
|  | S093 | NEG | NEG | 0 | - | N | Recovered/Non-Infected |
|  | S094 | NEG | NEG | 0 | - | N | Recovered/Non-Infected |
|  | S095 | NEG | NEG | 0 | 5 | N | Recovered/Non-Infected |
|  | S096 | NEG | NEG | 0 | - | N | Recovered/Non-Infected |
|  | S098 | NEG | NEG | 0 | - | N | Recovered/Non-Infected |
|  | S099 | NEG | NEG | 0 | - | Y | Recovered/Non-Infected |
|  | S100 | NEG | NEG | 0 | - | N | Recovered/Non-Infected |
|  | S101 | NEG | NEG | 0 | - | N | Recovered/Non-Infected |
|  | S102 | NEG | NEG | 0 | - | N | Recovered/Non-Infected |
|  | S103 | NEG | NEG | 0 | - | N | Recovered/Non-Infected |
|  | S104 | NEG | NEG | 0 | - | N | Recovered/Non-Infected |
|  | S105 | NEG | NEG | 0 | - | N | Recovered/Non-Infected |
|  | S106 | NEG | POS | 0 | - | N | Recovered/Non-Infected |
|  | S107 | NEG | NEG | 0 | - | N | Recovered/Non-Infected |
|  | S108 | NEG | NEG | 0 | - | N | Recovered/Non-Infected |
|  | S109 | NEG | NEG | 0 | - | N | Recovered/Non-Infected |
|  | S110 | NEG | NEG | 0 | - | N | Recovered/Non-Infected |
|  | S111 | NEG | NEG | 0 | - | N | Recovered/Non-Infected |
|  | S112 | NEG | POS | 0 | - | N | Recovered/Non-Infected |
|  | S113 | NEG | NEG | 0 | - | N | Recovered/Non-Infected |
|  | S116 | NEG | NEG | 0 | - | N | Recovered/Non-Infected |
|  | S123 | 35 | NEG | 0 | - | N | Recovered/Non-Infected |
|  | S124 | 36 | NEG | 0 | 48 | N | Recovered/Non-Infected |
|  | S125 | 39 | NEG | 0 | - | Y | Recovered/Non-Infected |

NEG= Negative result. POS= Positive result. Y = Presence of at least one symptom related to COVID-19 disease. N= Absence of symptoms related to COVID-19 disease. (-) = Not determined.

